# Evidence and characteristics of human-to-human transmission of SARS-CoV-2

**DOI:** 10.1101/2020.02.03.20019141

**Authors:** Min Kang, Jie Wu, Wenjun Ma, Jianfeng He, Jing Lu, Tao Liu, Baisheng Li, Shujiang Mei, Feng Ruan, Lifeng Lin, Lirong Zou, Changwen Ke, Haojie Zhong, Yingtao Zhang, Xuguang Chen, Zhe Liu, Qi Zhu, Jianpeng Xiao, Jianxiang Yu, Jianxiong Hu, Weilin Zeng, Xing Li, Yuhuang Liao, Xiujuan Tang, Songjian Xiao, Ying Wang, Yingchao Song, Xue Zhuang, Lijun Liang, Siqing Zeng, Guanhao He, Peng Lin, Huihong Deng, Tie Song

**Affiliations:** Guangdong Provincial Center for Disease Control and Prevention; Guangdong Provincial Institute of Public Health, Guangdong Provincial Center for Disease Control and Prevention; Shenzhen Center for Disease Control and Prevention; Zhuhai Center for Disease Control and Prevention; Shenzhen Nanshan Center for Disease Control and Prevention

## Abstract

**Background:** On December 31, 2019, an outbreak of COVID-19 in humans was reported in Wuhan, and then spread fast to other provinces, China. We analyzed data from field investigations and genetic sequencing to describe the evidence and characteristics of human-to-human transmission in Guangdong Province.

**Methods:** A confirmed COVID-19 case was defined if a suspected case was verified with positive of SARS-CoV-2 in throat swabs, nasal swabs, bronchoalveolar lavage fluid (BALF), or endotracheal aspirates by real-time reverse transcriptase polymerase chain reaction assay (RT-PCR) or genetic sequencing. Field investigations were conducted for each confirmed case. Clinical and demographic data of confirmed cases were collected from medical records. Exposure and travel history were obtained by interview.

**Results:** A total of 1,151 confirmed cases were identified as of February 10, 2020 in Guangdong Province, China. Of them, 697 (60.1%) cases were from 234 cluster infections. Two hundred and fourteen (18.6%) were secondary cases, in which 144 cases were from family cluster infections. With the epidemic continuing, although familial cluster events were dominated, community cluster events increased with a nosocomial event. The whole genomes within the same family cluster infections were identical, and presented a few unique single nucleotide variants (SNVs) compared with SARS-CoV-2 identified on December 2019 in Wuhan.

**Conclusions:** We observed evident human-to-human transmissions of SARS-CoV-2 in Guangdong, China. Although most of them were from family cluster infections, community and nosocomial infections were increasing. Our findings indicate that human-to-human transmission risks are transferring from family to community in Guangdong Province.

## Introduction

On December 31, 2019, Wuhan Municipal Health Commission reported 27 cases of pneumonia infection with unknown etiology in Wuhan, Hubei Province, China^1,2^. A novel coronavirus (SARS-CoV-2) was identified as the causative virus by Chinese authorities on January 7, 2020^3^. The common symptoms of the cases infected with SARS-CoV-2 include fatigue, fever, cough, shortness of breath and breathing difficulties. Severe infection can cause pneumonia, severe acute respiratory syndrome, kidney failure and even death^4^. As of February 11, 2020, 44,653 confirmed cases with 8,204 critical cases and 1,114 deaths were reported in 34 provinces, regions and municipal cities across China, with 33,366 in Hubei Province (19,558 cases in Wuhan), 1,219 in Guangdong Province, 1,135 in Henan Province, and 1,131 in Zhejiang Province. Additionally, 437 cases were reported in other 24 countries. Almost all provinces and regions in China have initiated the highest level of public health emergency response.

As an emerging infectious disease, the characteristics of COVID-19 such as natural host and reservoir, incubation period, serial interval, and infectivity of human-to-human transmission remain unknown. We described evidence and characteristics of human-to-human transmission of SARS-CoV-2 in Guangdong Province, which will be valuable to decision making on prevention and control strategies for community spreading.

## Methods

### Case definition and identification

The suspected and confirmed human infections with SARS-CoV-2 were defined based on the Diagnosis and Treatment Scheme of SARS-CoV-2 released by the National Health Commission of China (Supplementary appendix)^5^.

After the outbreak of COVID-19 in Wuhan announced on December 31, 2019, enhanced surveillance was initiated in Guangdong Province to early recognize suspected infections, especially among those who had history of travelling to Wuhan within 14 days. Once a suspected case was diagnosed, the Center for Diseases Control and Prevention (CDC) at prefecture level would conduct an initial field investigation, and collect respiratory specimens. The respiratory specimens were shipped to Guangdong Provincial CDC (GDCDC) for SARS-CoV-2 test by RT-PCR or genetic sequencing.

### Epidemiological data Collection

Demographic information, date of illness onset, and clinical outcomes of confirmed cases were collected from medical records. Through interviewing with confirmed cases or their relatives, we obtained exposure histories for each case during 14 days before the onset of illness including date, frequency, and patterns of exposures to wild animals.

### Metagenomic Sequencing

Clinical samples including bronchoalveolar lavage fluid (BALF), endotracheal aspirates, throat swabs, and nasal swabs were collected for suspected cases. Total RNAs were extracted and RT-PCR was firstly performed according to the standard method provided by the China CDC. Next, the positive samples with cycle threshold value lower than 35 were used for metagenomic sequencing to obtain the viral genomic sequences. Sample RNA was treated with TURBO DNase (Invitrogen, Cat. No. AM2239) and then purified by using Agencourt RNAClean XP beads (Beckman Cat. No. A63987). Libraries were prepared using the SMARTer Stranded Total RNA-Seq Kit v2 (Clontech, Cat. No. 634412) according to the manufacturer’s protocol. Pair-end sequencing was performed on Miseq (Illumina, USA).

### Statistical and Sequences analyses

We used descriptive statistics to summarize the epidemiological characteristics of confirmed cases. The sequencing data were quality controlled (QC) using fastp6 and then mapped to the human genome (hg38) using Bowtie27 to remove human related RNA reads. De novo assembling was performed using Spades8 and Megahit9 for each dataset. Assembled contigs were annotated using blastn10 using the complete nucleotide database. Twelve genome sequences generated in this study have been submitted to GISAID under the accession numbers EPI403932–EPI403937 and EPI406533–EPI406538. Maximum-likelihood (ML) trees of viral genome were estimated via IQ-Tree^11^ by integrating closely related sequences from SARs and SARs-like viruses from public database. Two SARS-CoV-2 genomes from introduction cases in Thailand were also included and data analysis was approved by the corresponding submitter. SNV (single nucleotide variant) sites were found through SNV-sites^12^. Phylogenetic trees were annotated and visualized with ggtree^13^.

### Ethics Approval

Data collection and analysis of cases and close contacts were determined by the National Health Commission of the People’s Republic of China to be part of a continuing public health outbreak investigation and were thus considered exempt from institutional review board approval.

## Results

### Description of the outbreak

As of February 10, 2020, a total of 1,151 cases were identified in 20 of 21 cities in Guangdong Province, with the majority of cases in Shenzhen (n=368) and Guangzhou(n=313). Most cases (81.4%, 937/1151) were imported from other Provinces, especially from Hubei Province (70.1%, 841/1,151). The peak of total cases was between January 23 and January 28, and so did for cluster cases and secondary cases (Figure 1). The average age of all cases was 44.5 years, and the percentage of males was 49.1% (565/1,151). Of total cases, 579 (50.3%) and 275 (23.9%) cases had history of travelling to Wuhan, and other cities in Hubei Provinces, respectively (Table 1). The average duration from onset of symptoms to diagnosis was 5.6±3.2 days.

**Table 1.** General characteristics of confirmed COVID-19 cases in Guangdong Province, China

| | Total (n, %) | Cluster cases<br>(n, %) | Non-cluster<br>cases<br>(n, %) | $\chi^2$ | P |
| --- | --- | --- | --- | --- | --- |
| **Sex** |  |  |  |  |  |
| Male | 565 (49.1) | 334 (47.9) | 231 (50.9) | 0.850 | 0.357 |
| Female | 586 (50.9) | 363 (52.1) | 223 (49.1) |  |  |
| **Age (years)** |  |  |  |  |  |
| <30 | 225 (19.5) | 141 (20.2) | 84 (18.5) | 9.295 | 0.098 |
| 30~ | 258 (22.4) | 151 (21.7) | 107 (23.6) |  |  |
| 40~ | 176 (15.3) | 99 (14.2) | 77 (17) |  |  |
| 50~ | 216 (18.8) | 121 (17.4) | 95 (20.9) |  |  |
| 60~ | 209 (18.2) | 138 (19.8) | 71 (15.6) |  |  |
| ≥70 | 67 (5.8) | 47 (6.7) | 20 (4.4) |  |  |
| **History of travelling to Wuhan**<br>**and other cities in Hubei** |  |  |  |  |  |
| Wuhan | 579 (50.3) | 320 (45.9) | 259 (57) | 33.66 | <0.001 |
| Other cities in Hubei | 275 (23.9) | 160 (23.0) | 115 (25.3) |  |  |
| No | 294 (25.5) | 217 (31.1) | 77 (17.0) |  |  |
| Unclear | 3 (0.3) | 0 (0.0) | 3 (0.7) |  |  |
| **Duration from onset to**<br>**diagnosis (days)** |  |  |  |  |  |
| <2 | 101 (8.8) | 81 (11.6) | 20 (4.4) | 43.26 | <0.001 |
| 2~ | 231 (20.1) | 160 (23) | 71 (15.6) |  |  |
| 4~ | 268 (23.3) | 157 (22.5) | 111 (24.4) |  |  |
| 6~ | 212 (18.4) | 132 (18.9) | 80 (17.6) |  |  |
| ≥8 | 290 (25.2) | 140 (20.1) | 150 (33) |  |  |
| Unclear | 49 (4.3) | 27 (3.9) | 22 (4.8) |  |  |
| **Severity of illness** |  |  |  |  |  |
| Mild | 993 (86.3) | 604 (86.7) | 392 (86.3) | 0.46 | 0.792 |
| Serious | 113 (9.8) | 66 (9.5) | 47 (10.4) |  |  |
| Critical | 42 (3.6) | 27 (3.9) | 15 (3.3) |  |  |

**Figure 1.**
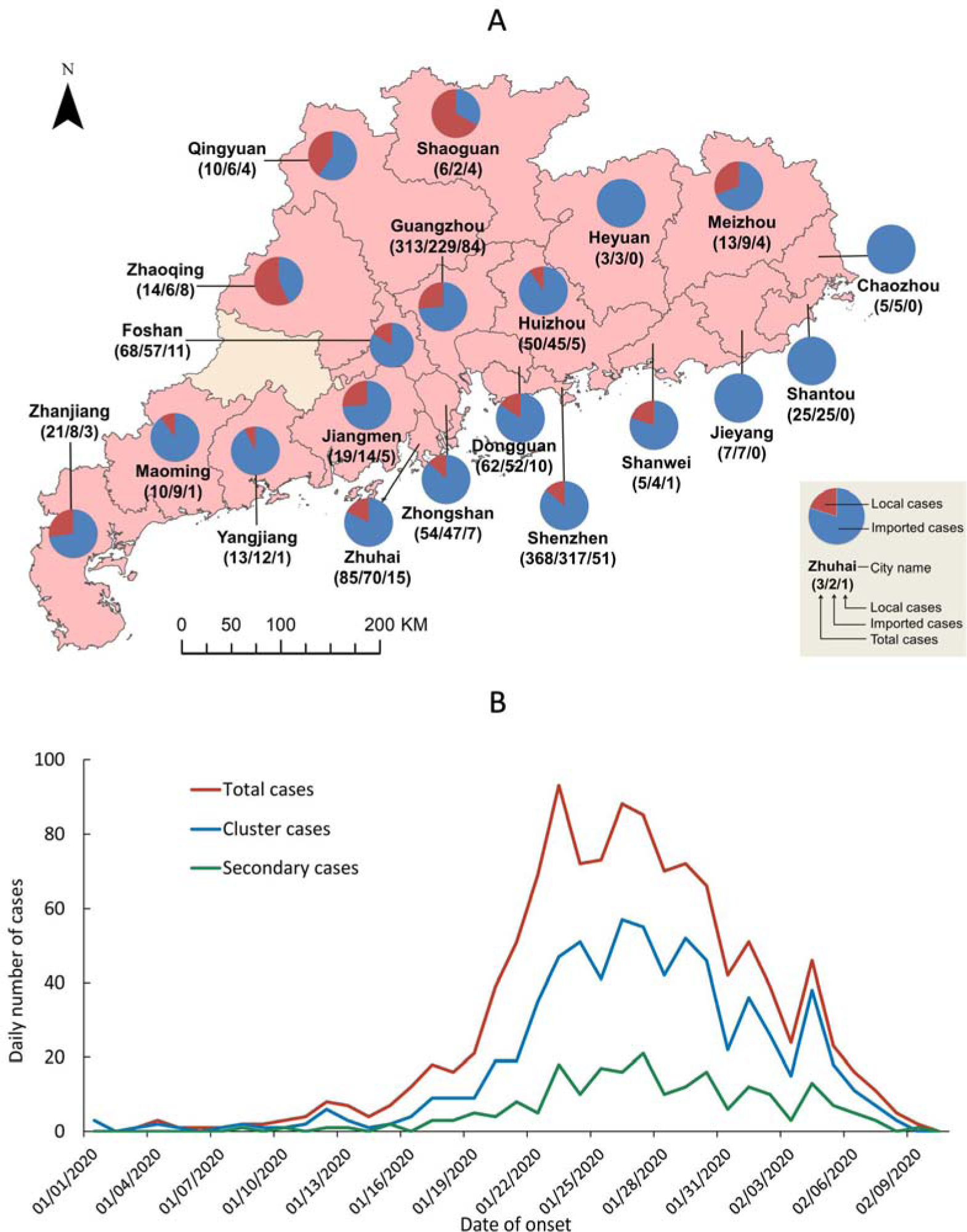
Spatio-temporal distribution of cases in Guangdong Province, China, by February 10, 2020. **Chart A:** The spatial distribution of all confirmed cases. **Chart B:** The temporal distribution three types of cases, and number of cluster infections.

Of total cases, 697 cases (60.1%) were from 234 cluster infection events with 537 cases in 195 family cluster events and 160 cases in community clusters. Figure 2 shows that most cluster infection events are familial clusters during the epidemic, and community cluster events increased since the end of January. More importantly, on February 4, 1 nosocomial cluster event (3 cases) were reported in a hospital. Two hundred and fourteen (18.6%) secondary cases were identified, where 144 cases were from family cluster infections, 56 from other cluster infections, and 14 from community infections.

**Figure 2.**
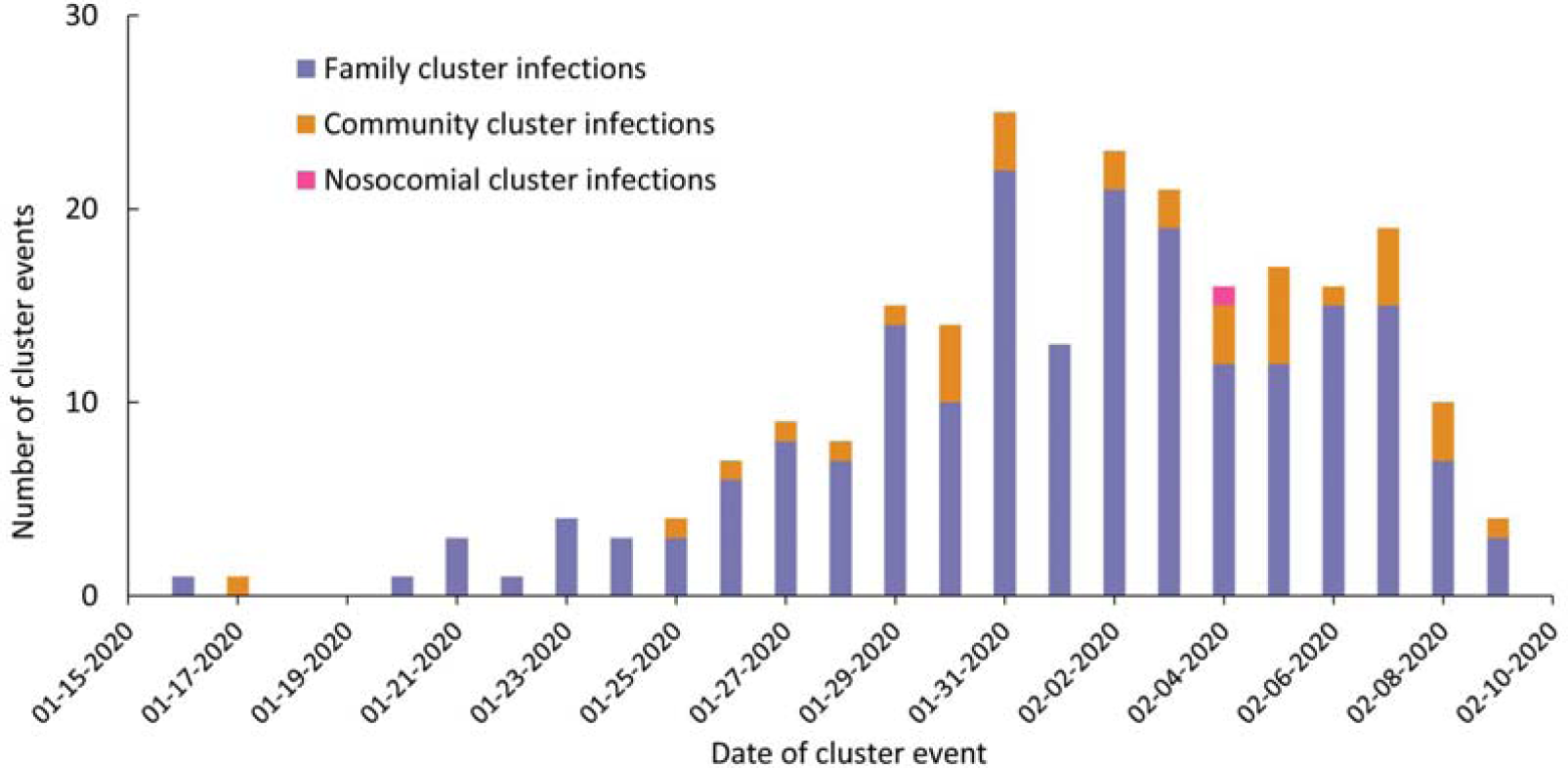
Time distribution of cluster infection events in Guangdong Province, by cluster types.

Here we extracted 13 family cluster infections as representatives of all family cluster cases to describe their characteristics, in which the epidemiological information of three family clusters were described in detail below. We also described two special events, one for community infection and another for nosocomial transmission.

### Characteristics of 13 family cluster infections

The 13 family cluster infections were reported from Shenzhen (n=4), Zhuhai (n=3), Guangzhou (n=2), Foshan (n=2), Shaoguan (n=1) and Yangjiang (n=1) (Figure 3). The most common symptoms in 37 cases included fever, fatigue, and cough. Of 37 cases, 3 and 10 cases were critically and severely ill, respectively. Three cases in family cluster E had only mild respiratory symptoms such as mild cough, and 2 cases in family cluster M did not report any symptoms. Thirty-four cases had no exposure to wild animals within 14 days prior to the onset of illness. Nine cases were identified as secondary cases (Figure S1 and Table S1).

**Figure 3.**
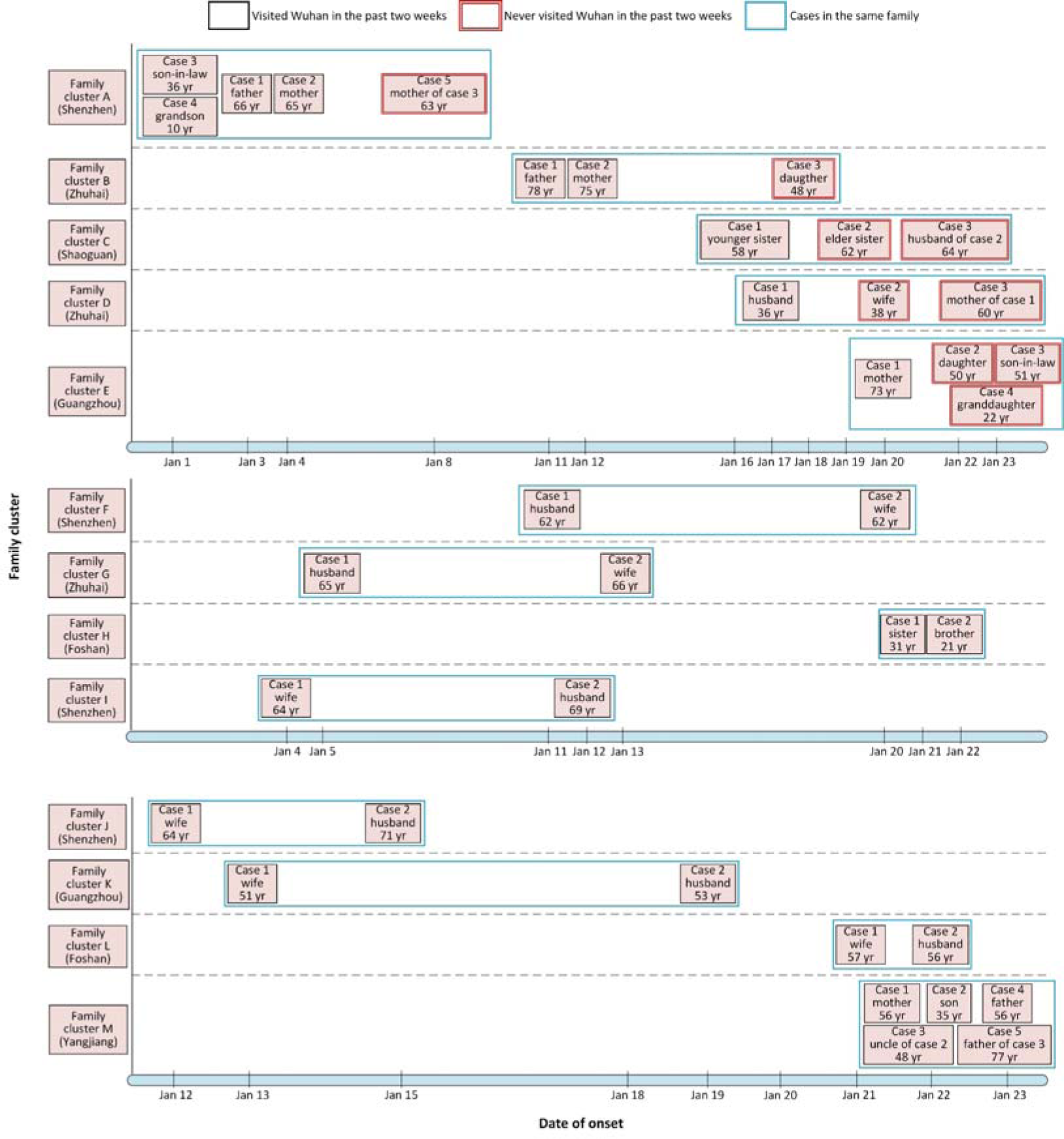
Onset date of 37 confirmed COVID-19 cases in 13 family cluster infections in Guangdong Province.

#### Family cluster A

This is the first family cluster infections identified in Guangdong Province. There were 5 cases in family cluster A. Case 1, Case 2, Case 3 and Case 4 stayed in Wuhan during December 29, 2019 to January 4, 2020. Case 1 had symptoms of cough, fatigue, fever and diarrhea on January 3, 2020 when they were in Wuhan. Case 2 (wife of Case 1) had fatigue and fever on January 4 after they returned Shenzhen. The couple visited two hospitals on January 6 and January 10 when they had fever (>38□) and chest radiologic changes. Their endotracheal aspirates were collected and tested by Center for diseases control and prevention (CDC) on January 13, and were identified positive infections of SARS-CoV-2. Case 3 (son-in-low of Case 1) and Case 4 (grandson of Case 1) had fever, stomachache, diarrhea and muscular soreness on January 1 in Wuhan, and visited a hospital on January 11 in Shenzhen. Their blood and throat swab samples were also identified positive with SARS-CoV-2. These four cases had no history of exposure to wild animals. Case 5 (mother of Case 3) did not visit Wuhan within 14 days before the onset of symptoms, but lived with Case 3 in an apartment in Shenzhen. On January 8, she had low fever and breathless. On January 14, she visited a hospital and was isolated for treatment. Her blood and throat swab samples were collected and tested by CDC on January 15, and identified positive of SARS-CoV-2.

#### Family cluster B

Three cases were identified in family cluster B in Zhuhai, including a father (Case 1), a mother (Case 2) and their daughter (Case 3). Case 1 and Case 2 resided in Wuhan, and visited on Case 3 in Zhuhai on January 11. On their trip to Zhuhai, Case 1 began to have symptom of fatigue. On January 12, Case 1 developed dry cough, fatigue, headache, eye pain and muscular soreness, and Case 2 had symptoms of dry cough, fatigue, headache. On January 15, Case 1 and Case 2 visited a hospital. Their throat swabs were collected and tested by CDC on January 17, and laboratory diagnosis showed positive of SARS-CoV-2. Case 3 had symptoms of fever, occasional itching and dry cough on January 17 without history of visiting Wuhan within 14 days prior to the onset of symptoms. She visited a hospital on January 18, and SARS-CoV-2 was identified in throat swab samples by CDC.

#### Family cluster D

Family cluster D in Zhuhai had three cases. Case 1 stayed at Wuhan from January 8 to 17, and returned Zhuhai on January 17. On the trip, he had symptom of muscular soreness. Case 2 (wife of Case 1) lived with Case 1 in Zhuhai and had cough on January 20 without history of visiting Wuhan within 14 days before the onset. She visited a hospital with Case 1 on January 20, and were transferred to another hospital on January 21 for treatment and isolation. Their nasal and throat swabs were tested and identified with positive of SARS-CoV-2 on January 22. Case3 (mother of Case 1) lived with Case1 and Case2 without history of visiting Wuhan in the past 14 days. Her nasal and throat swabs were also identified with positive of SARS-CoV-2 on January 22, when she had low fever and fatigue, and was isolated on January 23.

### Community infection event

Five cases were included in a community transmission event. Case 1 resided in Henan Province, China. On January 11, he and his wife (Case 2) came to Guangzhou to visit on their daughter (Case 3) by train which passed through Wuhan. Case 3 resided on the tenth floor of an apartment in Guangzhou. Case 1 and Case 3 had symptoms of fever on January 19, but they did not visit a hospital. Case 1 had symptoms of fever and dry cough on January 23. Case 1 and Case 2 visited a hospital on January 24 with Case 3, and their throat swabs were tested and identified with positive of SARS-CoV-2. They were isolated in a hospital for treatment. On January 16, Case 1 used the elevator in their apartment. Immediately after he got off the elevator, Case 4, who resided on the eleventh floor, entered the elevator, and touched herself mouth after she pressed the elevator button; she did not wear a mask. Case 4 did not meet Case 1, Case 2 and Case 3 before her onset of illness. On January 21, Case 4 had symptoms of fever, and visited a community hospital. On January 27, Case 4 and her husband (Case 5) visited another hospital, and their throat swabs were tested and identified with positive of SARS-CoV-2. Case 5 had no symptoms before he was diagnosed as COVID-19. Both Case 4 and Case 5 had no exposure to wild animals within 14 days prior to the onset of illness.

### Nosocomial infection event

Three COVID-19 cases were included in a hospital-associated transmission event. Case 1, who resided in Guangzhou, and visited her relatives in Hubei Province from January 12 to January 22. She had symptoms of fatigue and leg pain on January 27, and visited a hospital on January 28. She had fever when she was hospitalized. On February 2, she had chest radiologic changes, and her throat swabs were tested and identified with positive of SARS-CoV-2. Case 2 was admitted to the same room in the same hospital with Case 1 on December 31, 2019 because of backache. His white blood counts decreased on February 4, and he had severer cough on February 5 when he was identified with positive of SARS-CoV-2. Case 3 was a nurse who took care of Case 1 for several times, had symptoms of dry cough on February 4, and expectorated on February 5 when she was identified with positive of SARS-CoV-2. She had no exposure to wild animals within 14 days prior to the onset of illness, but did not strictly protect herself by using protective products when she nursed Case 1.

### Phylogenetic analyses

The phylogeny based on full-length of SARS-CoV-2 genomes (12 generated in this study) showed SARS-CoV-2 could be classified into SARS-like virus with the closest related strains as MG772934 and MG772933 which were collected from Rhinolophus sinicus in China on 2015 and 2017, respectively. SARS viruses shared nearly 97.5% nucleotide similarity with closely related SARS-like viruses from bats. By contrast, SARS-CoV-2 had lower nucleotide similarity with related SARS-like viruses (87.4%-87.5%), and a long internal branch was observed between SARS-CoV-2 and related SARS-like viruses (Figure 4A).

**Figure 4.**
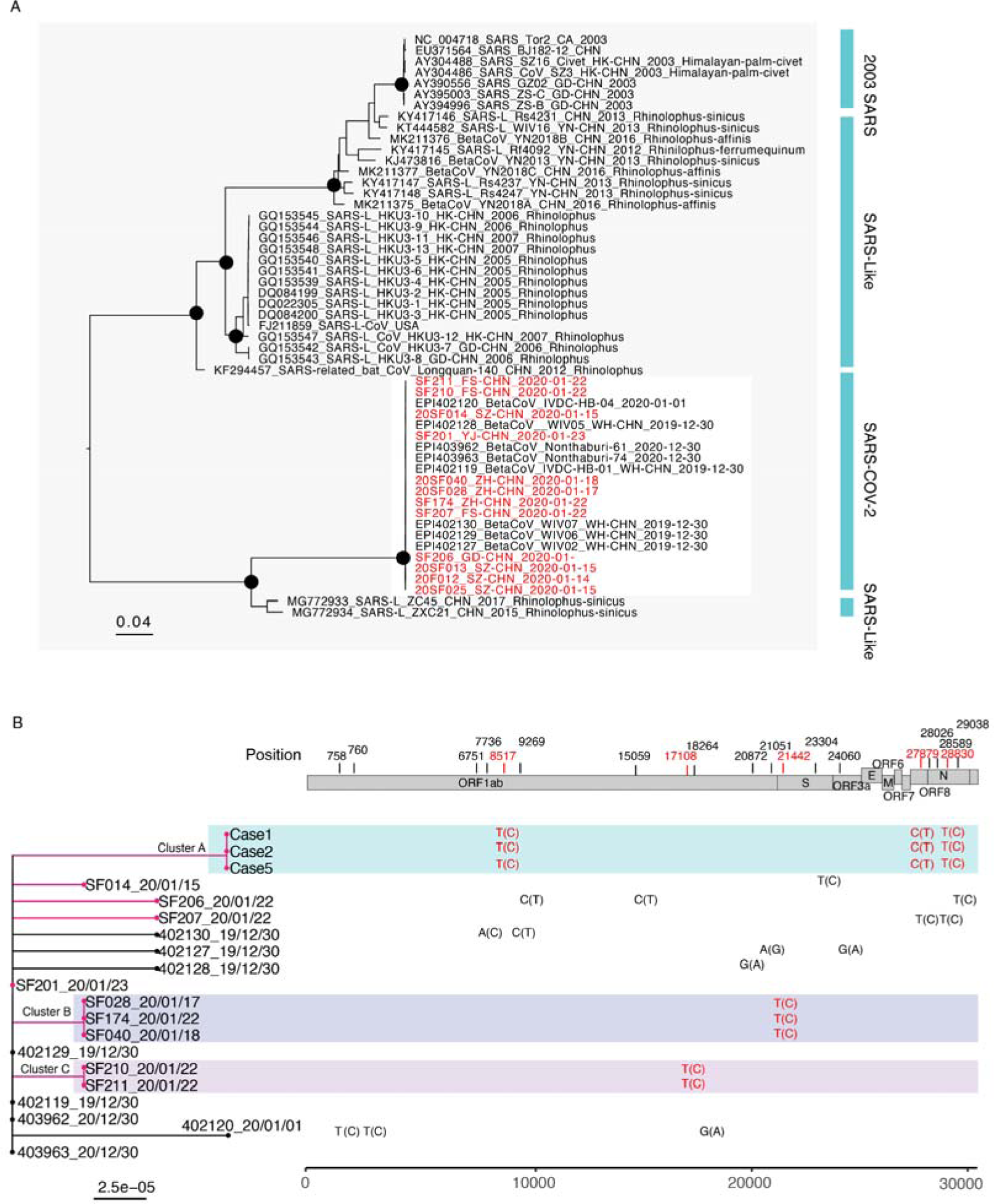
Phylogenetic analysis of the complete genome of SARS-CoV-2 identified in Guangdong and closely related viral genome from the public database. **Chart A**: Maximum-likelihood tree was constructed by including 12 new generated SARS-CoV-2 sequences in this study, 6 published SARS-CoV-2 sequences and sequences from closely related SARS-like viruses. Two SARS-CoV-2 sequences from Thailand were integrated into the analysis with the permission of the submitter. Black circles indicate bootstrap support >0.9 at the root node of selected clade. The cluster of SARS-CoV-2 was highlighted with red box. The sequences generated in this study were highlighted in red. **Chart B**: Phylogenetic relationship among SARS-CoV-2 and SNV (single nucleotide variant) sites were identified by aligning twenty complete genomes of SARS-CoV-2 collected between 30 December, 2019 and 23 January, 2020were found by the multiple genome sequences alignment. To ignore the sequences uncertainty at 5’ and 3’ terminal, the aligned sequences were started from position 266 to 29856 of SARS-CoV-2 genome (use the EPI402119 as the reference genome). The phylogeny branches of Guangdong SARS-CoV-2 sequences were colored in magenta and different family clusters were marked with different color boxes. Unique SNVs and their corresponding positions in reference viral genome identified in family cluster infections were shown on right of the phylogeny and the relative consensus sequences were shown in brackets. The nonsynonymous SNVs in cluster infections were marked with #.

To characterize the COVID-19 cases from family clusters and investigate potential genetic adaptation, we compared the twelve SARS-CoV-2 genome sequences generated between 14 January 2020 and 23 January 2020 in this study and all published SARS-CoV-2 ^14,15^. Sequences from the three family cluster infections fell into three corresponding genetic clusters and could be separated from the sequences of sporadic infections (Figure 4B). The viral genome sequences from cases within a family cluster were exactly the same. To find whether there were any genetic changes following the illness progress, we sequenced SARS-CoV-2 viruses from case 1 of cluster B on 17 January and 22 January, which was about 1 week and 2 weeks after the onset of illness, respectively. These two sequences were exactly the same suggesting the genetic stability of SARS-CoV-2 during the infection. Notably, the viral genome from family cluster A share three unique SNV sites which could not be found from other COVID-19 cases in public database. Among the three SNVs, one was predicted to cause amino acid changes (nonsynonymous variations) in ORF8 protein coding sequence led the amino acid change from Leu to Ser. One unique SNV site was identified in family cluster B and the change from C to T at position 21442 within the spike protein coding sequence leading to amino acid change from His to Thr. Only one synonymous variation was identified in viral genome sequences from family cluster C. Due to the increasing epidemic activity, we also investigated whether there were any parallel mutations, which was regard as an indicator of potential genetic adaptation, could be identified in recently identified sporadic infection cases. Three SARS-CoV-2 genetic sequences from sporadic cases on 22 January and 23 January showed there were no common SNV sites that could be identified in these cases and the SF201_20/01/23 SARS-CoV-2 collected on 23 January had exactly the same genetic sequence with viral sequences collected at the beginning of the epidemic.

## Discussion

Eighteen years after the first emergence of SARS-CoV in Guangdong at the end of 2002^16^, a great challenge presents by the emergence of a new SARS-like coronavirus (SARS-COV-2). The early epidemiological data suggested that most cases were caused by spill-over infections from nonhuman sources which is still unknown^17^. In our study, we described the characteristics of human-to-human transmissions of SARS-COV-2 using epidemiological and genetic evidence.

As of February 10, increasing secondary cases were identified in Guangdong Province. In all 214 secondary cases, none of them had history of residence or traveling to Wuhan within 14 days prior to onset of the illness. Our finding confirms the human-to-human transmission of SARS-COV-2 reported in a familial cluster infection in a previous study^18^, and further suggests that it has strong transmissibility between humans. We further observed several cases with mild symptoms or no symptoms, which indicates that current monitoring measures such as fever testing may not work effectively to identify these asymptomatic cases. With the epidemic ongoing, we found two cases infected by sharing elevator with an index case in a building, and increasing cases in gathering activities or communities. These findings suggest that the human-to-human transmission risks are transferring from familial cluster infections to community outbreak. More importantly, we observed a nosocomial transmission event in Guangdong Province, which was also reported in a hospital in Wuhan, where 57 patients including 40 health-care workers were infected^19^. “Super spreading events” may occur in some specific circumstances such as hospitals if the epidemic was not control on time. As the extended Chinese Spring Festival holiday ends, millions of people will travel back to Guangdong Province from regions across China, which will cause more cases to be imported to the Province. This will further increase the risk of community transmission. Therefore, we should persist in rigorous measures of cutting human-to-human transmission taken by the governments in communities especially in high risk places such as hospitals, factories and schools.

The evidence of strong human-to-human transmission raise much concern on 1) whether there are any genetic mutations associated with these cluster infections, and 2) are there any potential viral adaptations leading to the increasing epidemic activity? These questions, which are fundamental to the outbreak control, have not been adequately answered yet. In our study, the viruses sequenced from two secondary cases are the same with the viruses from corresponding index cases in their families, and these secondary cases had no history of traveling to Wuhan and no exposure to wild animals, which indicated that these viruses likely represent strains which can transmit from human to human. We also searched any genetic clues that might be associated with viral adaptation to a human transmission by comparing the viruses from family cluster infections with SARS-CoV-2 from the beginning of epidemic. Only three and one unique SNV sites are identified from viruses sequenced in family infection cluster A and B, respectively. These SNV sites are predicted to result in two synonymous change from Leu to Ser in Orf 8 and from His to Thr in spike protein, respectively. It is still unknown whether these amino acid changes provide or increase the virus capability of human transmission. However, the previous study from SARS suggests Orf8 is either noncoding or coding for a functionally unimportant putative protein^20^. As a result, few nonsynonymous changes detected between COVID-19 cases from secondary cases in Guangdong and early COVID-19 cases in Wuhan indicates SARS-CoV-2 could already transmit among humans at the beginning of the epidemic when it was still regarded as spill-over infections.

In this circumstance, more human to human transmissions should occur at the end of December 2019 than we previously expected. Another reason that we believe SARS-CoV-2 may be already well established in humans since its first identification is that the SARS-CoV-2 we identified on January, 23, 2020 were exactly the same with viruses identified in Wuhan on 30 December 2019. Together with the SARS-CoV-2 genome sequences generated on 15 and 22 January, we did not find any clues of adaptations within the genome of SARS-CoV-2 accompany with the increasing epidemic. These data suggest the genetic stability of SARS-CoV-2 during the epidemic, which is a contrast to our previous findings on SARS viruses for which the strong positive selection on spike protein coding sequence and deletions in ORF8 gene are observed at the early phase of the outbreak^20^.

Our findings indicate that with the epidemic ongoing, prevention measures of containing person-to-person transmission in communities are urgently enhanced. Persistent surveillance for COVID-19 in communities, early detection and isolation of contacts of confirmed cases, and virologic analyses to assess genetic changes that might suggest increased transmissibility among humans are all critical to inform prevention and control efforts and assessing the global pandemic potential of COVID-19.

## Data Availability

Data are applied through the corresponding author.

## Acknowledgements

We thank all the medical and nursing staff who assisted in the care of patients; the members from health department and CDC in Guangdong Province for their contribution in data collection, COVID-19 control and prevention; Dr. Pilailuk Okada and his colleagues for sharing their sequence data. This work was supported by National Key Research and Development Program of China (2018YFA0606200, 2018YFA0606202), the Science and Technology Program of Guangdong Province (2018B020207006, 2019B020208005, 2019B111103001), Guangzhou Science and technology Plan Project (201804010383).

